# Stroke recovery beyond initial severity: The complementary roles of brain structure and brain function in acute stroke

**DOI:** 10.1101/2025.08.24.25333888

**Authors:** Catharina Zich, Lydia C Mardell, Andrew J Quinn, Sven Bestmann, Nick S Ward

## Abstract

Stroke frequently results in long-term upper limb (UL) motor impairments, limiting independence and quality of life. Accurate prediction of recovery trajectories is essential for personalizing rehabilitation strategies. While structural brain metrics such as corticospinal tract (CST) integrity have been widely studied, they incompletely explain motor outcome variability. Functional brain activity, quantified by sensorimotor activity in the beta (β) frequency range has emerged as a promising biomarker of motor system integrity and plasticity potential.

This study assessed in 30 acute stroke survivors and 26 healthy controls how combining functional and structural metrics of brain function relates to initial motor severity and subsequent recovery, using clinical MRI/CT and electroencephalography during passive finger movement and rest.

Structurally, grey and white matter damage were associated with initial severity. No associations with recovery were found for structural metrics alone. Functionally, β-activity in response to passive movement, and resting state activity were related to recovery, independent of initial impairment. Multivariate regression revealed that combining initial severity, structural information (CST damage) and brain function (sensorimotor β activity) provided the most accurate prediction of both global and UL-specific recovery (R² = 80.1% and 74.3%, respectively).

These findings underscore the importance of integrating functional and structural neural markers for improved stroke outcome prediction.

## 1. Introduction

Stroke is a leading cause of long-term disability, with one in four people affected in their lifetime. Among survivors, about 25% remain moderately to severely disabled after ten years (Feigin et al., 2023). Improving long-term outcomes requires an understanding of the mechanisms of both initial impairment and subsequent recovery trajectories (Ward, 2017). Here, we focus on upper limb (UL) impairment, which is common and often limits daily independence and quality of life (Broeks et al., 1999). Long-term UL motor outcomes after stroke are related to initial impairment (Coupar et al., 2012), which itself is related to the characteristics of damage to brain structures. For example, the integrity of descending white matter (WM) pathways, most notably the corticospinal tract (CST), is associated with the severity of initial UL impairment in the acute stage after stroke (Aikio et al., 2021; Boccuni et al., 2019; Maraka et al., 2014; Saltão da Silva et al., 2022; Swayne et al., 2008).

However, it is still the case that stroke survivors with the same level of initial UL impairment can have very different long-term UL outcomes, especially in those with more severe initial UL impairment (Bonkhoff et al., 2022; Prabhakaran et al., 2008; Winters et al., 2015; Zich et al., 2025). These differences in long-term UL outcomes can be partly explained by different levels of post-stroke CST integrity (Byblow et al., 2015; Feng et al., 2015; Puig et al., 2017; Saltão da Silva et al., 2022; Spampinato et al., 2017) but substantial unexplained variability in long-term UL outcomes remains. This raises the question whether the mechanisms of initial impairment and the mechanisms of subsequent recovery are different and potentially independent (Ward, 2017; Zich et al., 2025). The International Stroke Recovery and Rehabilitation Alliance consensus paper suggested that the mechanisms of recovery might be more closely related to post-stroke brain function, highlighting several potential biomarkers, including cortical oscillatory activity as measured with electroencephalography (EEG) or magnetoencephalography (MEG) (Boyd et al., 2017). Identifying brain function biomarkers of the recovery process itself, independent from the mechanisms of initial impairment would be an important finding since brain function is more amenable to therapeutic intervention for enhancing recovery than brain structure (Ward, 2017).

Sensorimotor beta (β) activity (13–30 Hz) is a promising biomarker of motor function. Movement is accompanied by β-suppression (event-related desynchronisation, ERD) reflecting sensorimotor cortex activation, followed by a rebound (event-related synchronisation, ERS) linked to cortical inhibition or deactivation (Chen & Hallett, 1999; Franzkowiak et al., 2010; Pfurtscheller, 1992). This ERD-ERS pattern is robust and reproducible (Espenhahn et al., 2019; Illman et al., 2021). β-activity is driven by GABA-A–mediated inhibitory interneuron activity (Hall et al., 2010; Jensen et al., 2005; Yamawaki et al., 2008) and so reflect shifts in the cortical excitatory-inhibitory balance that could mediate experience-dependent plasticity (Ward, 2017). Indeed, sensorimotor cortex β-activity reflects motor skill learning ability in healthy controls (Haar & Faisal, 2020; Tan et al., 2014; Torrecillos et al., 2018; M. Wu et al., 2025) and stroke survivors (Espenhahn et al., 2020), but can be reduced after stroke (Laaksonen et al., 2012; Parkkonen et al., 2017, 2018; Tang et al., 2020; Zich et al., 2025).

In this study, we used routinely collected stroke service data, as well as bedside observations, to support future implementation in clinical practice. We hypothesised that (i) initial UL impairment is most closely related to measures of brain structure; but that (ii) subsequent UL recovery is most closely related to measures of brain function (sensorimotor β-activity); and that (iii) measures of brain function would account for some variability in subsequent UL recovery, independent of initial impairment and brain structure, highlighting early brain function as a potential therapeutic target for enhancing recovery.

## 2. Materials and methods

We obtained EEG data during a passive movement task and at rest during a single session from stroke survivors in the acute stage post-stroke and healthy controls. EEG data were complemented by clinical data and brain structural data (i.e., routine magnetic resonance imaging [MRI] and computed tomography [CT]).

### 2.1 Experimental design

#### 2.1.1 Ethical approval

The Research Ethics Committee of the University College London and the NHS Research Ethics Committee (London - Surrey Research Ethics Committee) approved the study protocol (20/LO/0520), and all subjects provided written informed consent.

#### 2.1.2 Subjects

Acute stroke survivors (Bernhardt et al., 2017) with first-ever stroke, aged 18 years and older, were recruited from the Hyper-acute stroke unit and acute stroke unit at The National Hospital for Neurology and Neurosurgery (NHNN), Queen Square. Exclusion criteria comprised: medically unstable, requiring neurosurgical intervention, severe language/cognitive deficits preventing consent, inability to understand English, or sign their name.

#### 2.1.3 Clinical data

Initial global severity was measured using the NIH Stroke Scale (NIHSS), with UL-specific severity based on its UL sub-scores. The NIHSS, which reflects severity of neurological impairment at stroke onset, was administered at admission (M = 6.6, SD = 6.0; range: 0–20 days post-stroke).

Global subsequent recovery was assessed using the total score of the Stroke Impact Scale (SIS) at six months post-stroke, adjusted for initial NIHSS score. UL-specific recovery was measured by the UL subscale of the SIS, also corrected for the initial UL NIHSS sub-score. The SIS was collected at an average of 6.2 months post-stroke (SD = 0.3; range: 5.8–7.1 months) via online questionnaire, paper form, or phone interview, depending on patient preference.

### 2.2 EEG data

#### 2.2.1 EEG data acquisition

EEG data were recorded with sintered Ag/AgCl electrodes from 30 scalp sites using electrode caps (Easycap, Herrsching, Germany), with AFz as the ground and FCz as the reference. Data were acquired with an amplitude resolution of 0.1 μV and a sampling rate of 500 Hz, using online analogue filter settings of 0.02 to 250 Hz, via a wireless amplifier (SmartingPRO, mBrainTrain, Belgrade, Serbia) attached to the back of the EEG cap. Data were transmitted via Bluetooth 5.0 to a laptop (Dell Latitude 7390) via a Bluetooth dongle (VCK dongle). No data were lost during transmission. Impedances were checked at the beginning (Smarting Streamer software v4.1; mBrainTrain, Belgrade, Serbia) and recorded throughout the experiment with a sampling rate of 125 Hz. Impedances were on average 17.4 kΩ (*SD* = 9.4 kΩ) in stroke survivors and 15.5 kΩ (*SD* = 5.9 kΩ) in HC.

Patient data were recorded on five active hospital wards of the hyper-acute stroke unit and acute stroke unit at The National Hospital for Neurology and Neurosurgery (NHNN), Queen Square. Stroke survivors were, depending on their clinical condition and preference, either in a sitting or supine position. Data from HCs were recorded in a side room of the same building while they were seated.

#### 2.2.2 Passive movement task and resting state

Data were collected during passive movement of the left and right index fingers using a block design. We used a passive movement as this provides access to sensorimotor β-activity, unconfounded by residual movement. The subject’s index finger was positioned above a keyboard key (Ecarke OSU Keypad 4-Key Gaming Keyboard) and held in place using a Velcro sleeve. The experimenter moved the key down and immediately up roughly every 5 s (Stroke survivors: *M* = 5.1 s, *SD* = 0.2 s; HC *M* = 5.1 s, *SD* = 0.1 s) for 6 min (Stroke survivors: *M* = 67.3, *SD* = 3.1, range 61-74 trials; HC *M* = 67.9, *SD* = 1.4, range 64-70 trials). Participants were instructed to relax, not to move their head or fingers, and not to pay attention to the stimuli. Moreover, data were collected during rest with eyes open (Stroke survivors: *M* = 7.0 min, SD = 0.4 min; HC *M* = 7.0 min, *SD* = 0.1min). Due to time constraints resting state data could not be obtained in nine stroke survivors.

#### 2.2.3 EEG data pre-processing

EEG data were preprocessed with the EEGLAB toolbox version v2021.1 (Delorme & Makeig, 2004) for MATLAB (version 2022a; MathWorks, Natick, MA, USA). First, to suppress high frequencies, data were low-pass filtered (35 Hz, FIR, hamming window, filter order: 166). Data were downsampled to 250 Hz to reduce computational demands and disk space consumed, and high-pass filtered at (1 Hz, FIR, hamming window, filter order: 826) to suppress very slow trends in the data. Edge artefacts were removed by discarding the first and last 5 s of each recording. Identification of improbable channels was conducted using the EEGLAB extension trimOutlier (https://sccn.ucsd.edu/wiki/EEGLAB_Extensions) with an upper and lower boundary of two standard deviations of the mean standard deviation across all channels (Stroke survivors: *M* = 1.6, *SD* = 0.6, range 0-3 channels; HC *M* = 0.5, *SD* = 0.9, range 0-3 channels). Channels exceeding this threshold were replaced by spherical interpolation. Two copies of the dataset were created. One copy was cleaned using Artifact Subspace Reconstruction (ASR, as implemented in EEGLAB plugin clean_rawdata(), (Blum et al., 2019; T. Mullen et al., 2013; T. R. Mullen et al., 2015)) aiming to remove non-stereotypical artefacts to improve Independent Components Analysis (ICA). We used temporal ICA across the channels using the runica algorithm (Makeig et al., 1995) to estimate 30 independent components. Components reflecting line noise, eye, muscle and heart activity were identified using the EEGLAB extension ICLabel (Pion-Tonachini et al., 2019) and controlled by visual inspection. Components flagged as artefactual (Stroke survivors: *M*=6, *SD*=2.9, range 4-10 components; HC *M*=5.9, *SD*=2.3, range 4-11 components) were regressed out of the second copy created after spherical interpolation.

For the resting state data, the power spectral density (PSD) estimate was computed with a 2 second window with a 1 second overlap. The spectra were normalised to represent relative power by dividing each bin in the spectrum by the total power across all bins. For each participant, this 2-dimensional array of the spectrum for each channel (frequencies × sensors) was computed and used in the group analysis.

Task data were segmented from -1.5 to 3.5 s relative to the button press. Artefactual epochs as indicated by the joint probability (EEGLAB function pop_jointprob.m, *SD* = 3) were discarded from further analyses (Stroke survivors: *M* = 11.1%, *SD* = 3.4%, range 7.3-18.9%; HC *M* = 10.3%, *SD* = 3.4%, range 5.9-24.8%). Time-frequency representations of individual trials were calculated using the Fieldtrip functions (www.fieldtriptoolbox.org; (Oostenveld et al., 2010)) integrated in SPM12 using Morlet wavelet analysis with a wavelet width of 9. Power values were baseline corrected (-1 to -0.2 s). Data were averaged across the β-(13-30 Hz) frequency range, resulting in two-dimensional data (time x space).

### 2.3 MRI and CT data

#### 2.3.1 MRI and CT data acquisition

CT (N = 28) and MRI (N = 27) data were acquired as part of the hospital routine. T1 and T2 sequences (TR = 4000 ms; TE = 101 ms; voxel size = 1.59 ×1.59 ×6.5 mm^3^; matrix = 172 × 172, 23 slices; flip angle = 90 deg) were obtained using a 1.5T Siemens Avanto MRI scanner (Siemens Healthineers, Germany). CT sequences (matrix = 512 × 512; with 34, 108, 112, 217 slices) were obtained using a Siemens GoAll or Siemens XCite CT scanner.

#### 2.3.2 Lesion mapping and quantification of lesion volume, GM damage and WM damage

Stroke lesions were demarcated using the semi-automated segmentation algorithm *Clusterize* (https://www.medizin.uni-tuebingen.de/de/das-klinikum/einrichtungen/kliniken/kinderklinik/kinderheilkunde-iii/forschung-iii/software) applied to axial T2, T1 or CT images. Agreement between a manual segmentation and the semi-automated lesion maps obtained with *Clusterize* has been previously demonstrated (de Haan et al., 2015; Ito et al., 2019; Wilke et al., 2011). The resulting lesions were manually verified and, if necessary, manually corrected. Lesion maps were smoothed using a 2 mm full-width half maximum (FWHM) Gaussian kernel. Lesions were normalised to standard MNI space and left hemispheric lesions were flipped.

We calculated the lesion volume, GM damage and WM damage using the *Lesion Quantification Toolkit* (https://wustl.app.box.com/v/LesionQuantificationToolkit)(Griffis et al., 2021). Gray matter (GM) damage is quantified as the percentage of voxels within each region of the automated anatomical labeling (AAL) atlas (Tzourio-Mazoyer et al., 2002) that overlapped with the lesion. We obtained GM damage for each region separately as well as total GM damage (i.e., mean across all regions). White matter (WM) damage is quantified as the percentage of streamlines of the HCP842 tractography atlas (Yeh et al., 2018) that intersect the lesion. We obtained WM damage for each tract separately as well as total WM damage (i.e., mean across all tracts).

### 2.4 Statistical analysis

Brain functional data were analysed using a General Linear Model (GLM) framework as implemented in glmtools (https://pypi.org/project /glmtools/; (Quinn et al., 2024)). Two GLMs (task, rest) were constructed to compare group averages, each with three regressors: two categorical predictors using dummy coding for participant group (healthy/acute), and one continuous covariate representing z-transformed participant age. Parameters were estimated using the Moore-Penrose Pseudo-Inverse for each frequency–sensor pair in the resting-state data, resulting in a spectrum of parameters. For the task data, parameters were estimated for each time–sensor pair, yielding a time course of parameters. A contrast of parameter estimates was defined to test for differences in the parameters between the healthy and acute groups. The resulting contrast estimates (COPEs) and their associated standard errors are used to compute a t-test for this contrast.

Four additional parametric models were constructed to assess correlations with both initial impairment and subsequent recovery, while accounting for initial impairment, in the patient group for both task and rest data. Specifically, two GLMs were developed to examine correlations with initial impairment (NIHSS) for task and rest data. Each model included one constant regressor and one parametric regressor, comprising the z-transformed NIHSS values for each participant. Additionally, two further models were constructed to assess subsequent recovery (SIS), while controlling for initial impairment (NIHSS), again for both task and rest data. These models contained a single constant regressor and two parametric regressors containing the z-transformed values for SIS and NIHSS for each participant. Parameters were estimated using the Moore-Penrose Pseudo-Inverse and a contrast of parameter estimates defined to test for differences between the parameters of each variable and zero. The resulting contrast estimates (COPEs) and their associated standard errors are used to compute a t-test for this contrast.

Null-hypothesis testing for structural and functional data was conducted using cluster based non-parametric permutation tests (Maris & Oostenveld, 2007; Nichols & Holmes, 2001). This approach makes minimal assumptions, addresses the issue of multiple comparisons, and is suitable when parametric assumptions are not met (Nichols & Holmes, 2001). Row-shuffle permutations were used to compare subgroup differences and test relationships between variables, while sign-flipping permutations were used to test whether a measurement deviated from zero. A null distribution of cluster statistics was generated by permuting the data and calculating the sum of *t*-values within the largest contiguous cluster exceeding a cluster-forming threshold of *t* = 3.5. Observed cluster statistics were compared to this null distribution and considered significant if they exceeded a predefined critical value. In this study, the null distribution was constructed using 5,000 permutations, with the 95th percentile used as the significance threshold.

Prediction analysis was conducted using R (version 4.0.2) and predictors were standardised using z-transformation. First, a linear regression with initial severity as predictor and subsequent recovery as dependent variable was performed. This initial model was compared to models in which brain structure and brain function were added separately as predictors. These models were, in turn, compared to a model comprising initial severity, brain structure, and brain function as predictors. The results were confirmed by forward stepwise linear regression. To this end, at each step, predictors were included when *p* < 0.15 (Wald test) and removed when *p* >= 0.15 (Wald test). Predictors showing high collinearity (variance inflation factor (VIF) > 2.5) were re-assessed. *R^2^* and *RMSE* were used to quantify the models’ overall ability to predict recovery. Moreover, the relationship between each predictor and the dependent variable was described by the regression coefficients.

### 2.5 Data availability

All data produced in the present study are available upon reasonable request to the authors.

## 3. Results

### 3.1 Participants

375 acute stroke survivors were screened within 7 days of stroke, of which 32 were recruited (**Fig. 1a**). Two datasets were excluded: one participant aborted the session, and one dataset was corrupted by an unspecific artefact, resulting in 30 datasets for the main analysis (4 females, age: *M* = 64.7; *SD* = 13.5; range 30.3-94.4 years, **Table 1**, **Fig 1b_i_**). At the time of testing the time since stroke was *M* = 3.5; *SD* = 1.8; range 1-7 days (**Fig 1b_ii_**). Three stroke survivors did not complete the SIS follow-up, resulting in 27 datasets for the prediction analysis. Data from 26 HC were collected (9 females, age: *M* = 56.5; *SD* = 11.1; range 37.7-85.2 years, **Fig 1b_i_**).

**Fig. 1.**
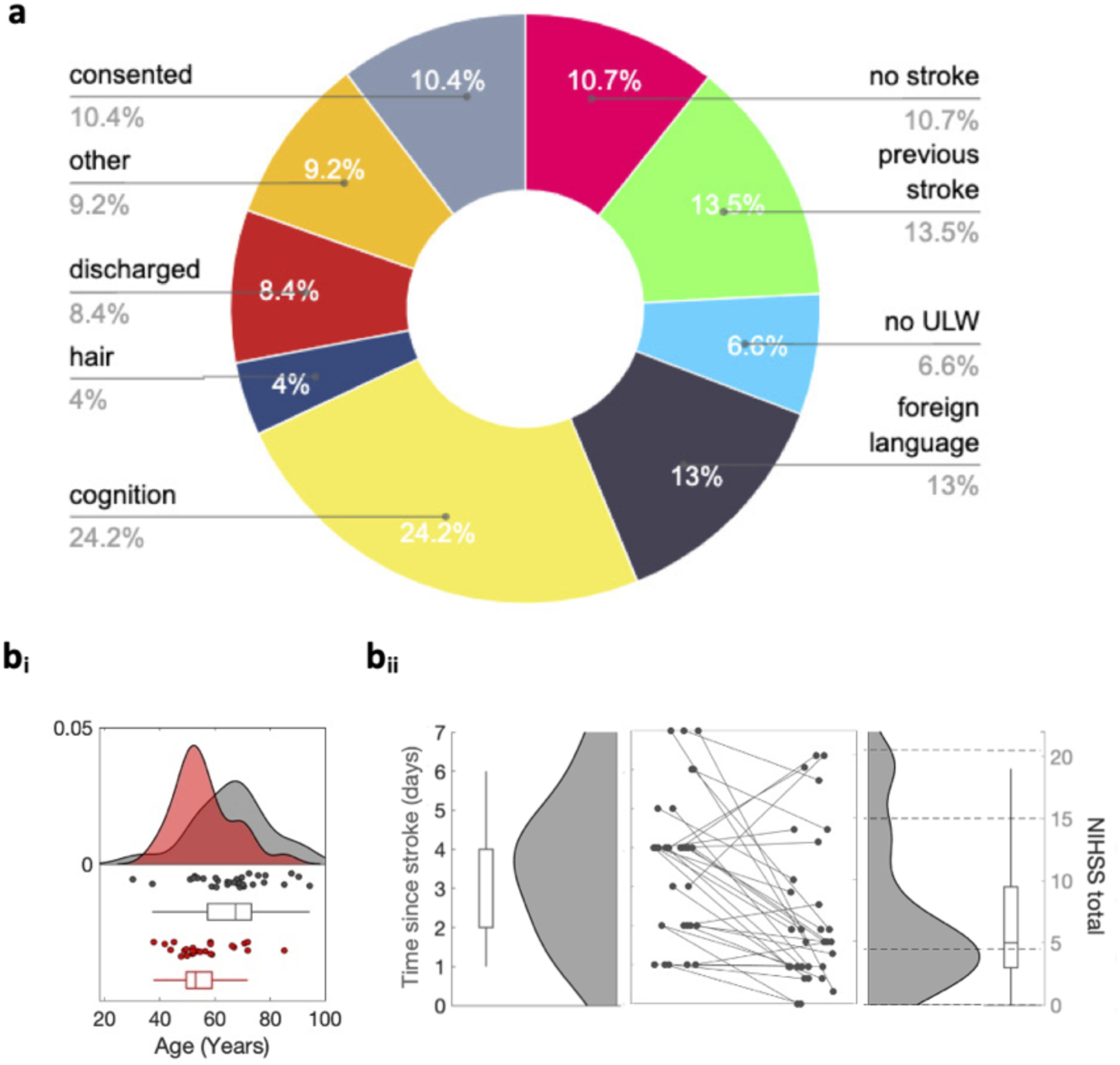
Recruitment chart and selected Demographics. **a**) Inclusion and exclusion criteria for 375 screened patients. **b_i_**) Age for healthy controls (red, N = 26) and stroke survivors (grey, N = 32). Data are shown as probability function (top), individual subjects, and boxplot (bottom). **b_ii_**) Time since the stroke and NIHSS total for stroke survivors. Data are shown as boxplot, probability function, and individual subjects (from the outside to the inside).

**Table 1.** Clinical details of the stroke survivors.

| ID | Age | Gender | Days | Type | Affected Hand | Location/description | NIHSS |
| --- | --- | --- | --- | --- | --- | --- | --- |
| 1 | 66-70 | M | 1 | I | R | Pons | 3 |
| 2 | 71-75 | M | 1 | H | L | BG | 3 |
| 3 | 66-70 | M | 1 | I | R | Precentral gyrus | 0 |
| 4 | 36-40 | M | 6 | H | R | Pons | 6 |
| 5 | 51-55 | M | 2 | I | R | Striato-capsular | 4 |
| 6 | 51-55 | M | 7 | H | L | Parietoccipital | 1 |
| 7 | 76-80 | F | 2 | I | L | Precentral gyrus | 2 |
| 8 | 66-70 | M | 4 | I | R | Corona Radiata | 3 |
| 9 | 66-70 | M | 2 | I | R | PCA | 3 |
| 10 | 86-90 | F | 2 | I | L | Precentral gyrus | 2 |
| 11 | 26-30 | M | 4 | I | R | Precentral gyrus | 6 |
| 12 | 51-55 | M | 2 | I | L | PICA | 0 |
| 13 | 56-60 | M | 5 | I | L | BG, Corona Radiata | 3 |
| 14 | 71-75 | F | 4 | I | L | Perirolandic region, corona radiata | 9 |
| 15 | 51-55 | M | 4 | H | R | BG | 13 |
| 16 | 56-60 | M | 4 | I | L | Posterior lentiform | 5 |
| 17 | 66-70 | M | 4 | I | L | Hemipons | 5 |
| 18 | 71-75 | M | 3 | I | R | M1, BG | 20 |
| 19 | 61-65 | M | 3 | I | L | MCA, M1 | 2 |
| 20 | 61-65 | M | 4 | I | L |  | 19 |
| 21 | 61-65 | M | 4 | I | L | BG | 6 |
| 22 | 81-85 | M | 2 | I | L | Frontal & parietal lobe | 5 |
| 23 | 66-70 | M | 4 | H | L | temporo-occipital lobe | 3 |
| 24 | 91-95 | M | 4 | I | L | MCA, posterior-inferior frontal lobe & insula | 6 |
| 25 | 66-70 | M | 1 | I | R | MCA, S2 | 5 |
| 26 | 51-55 | M | 7 | H | L | BG | 18 |
| 27 | 61-65 | M | 7 | I | L | L PICA, R PCA | 5 |
| 28 | 51-55 | F | 6 | H | R | Thalamus | 14 |
| 29 | 66-70 | M | 2 | I | R | Striatocapsular | 8 |
| 30 | 56-60 | M | 4 | I | L | Large MCA | 20 |
| Stroke (N=30) | 64.7 (30.3-94.4) | 26M/4F | 3.5 (1-7) | 23I/7H | 18L/12R |  | 6.6 (0-20) |
| Controls (N=26) | 56.5 (37.7-85.2) | 17M/9F | - | - | - | - | - |

M = male; F = female; I = ischemic; H = haemorrhagic; R = right; L = left; NIHSS = National Institutes of Health Stroke Scale.

### 3.2 Stroke-related changes in brain structure and function

First, we examined stroke-related changes in brain structure. *Lesion volume:* Stroke-related damage to brain structure particularly involved subcortical grey and WM structures in the middle cerebral artery (MCA) territory (**Fig. 2a_i_**) (mean lesion volume = 16.92 ml, *SD* = 18.29 ml, range 1.01-63.56 ml, **Fig. 2a_ii_**). *Grey matter:* GM damage most commonly involved the pallidum, putamen and amygdala (**Fig. 2b_i_**, **Table 2**) (mean total GM damage = 0.97%, *SD* = 1.20%, range 0.03-4.69%, **Fig. 2b_ii_**). *White matter:* WM damage most commonly involved corticospinal, parietopontine, and frontopontine tracts (**Fig. 2c_i_**, **Table 3**) (mean total WM damage = 7.78%, *SD* = 7.36%, range 0.34-34.90%, **Fig. 2c_iii_**).

**Fig. 2.**
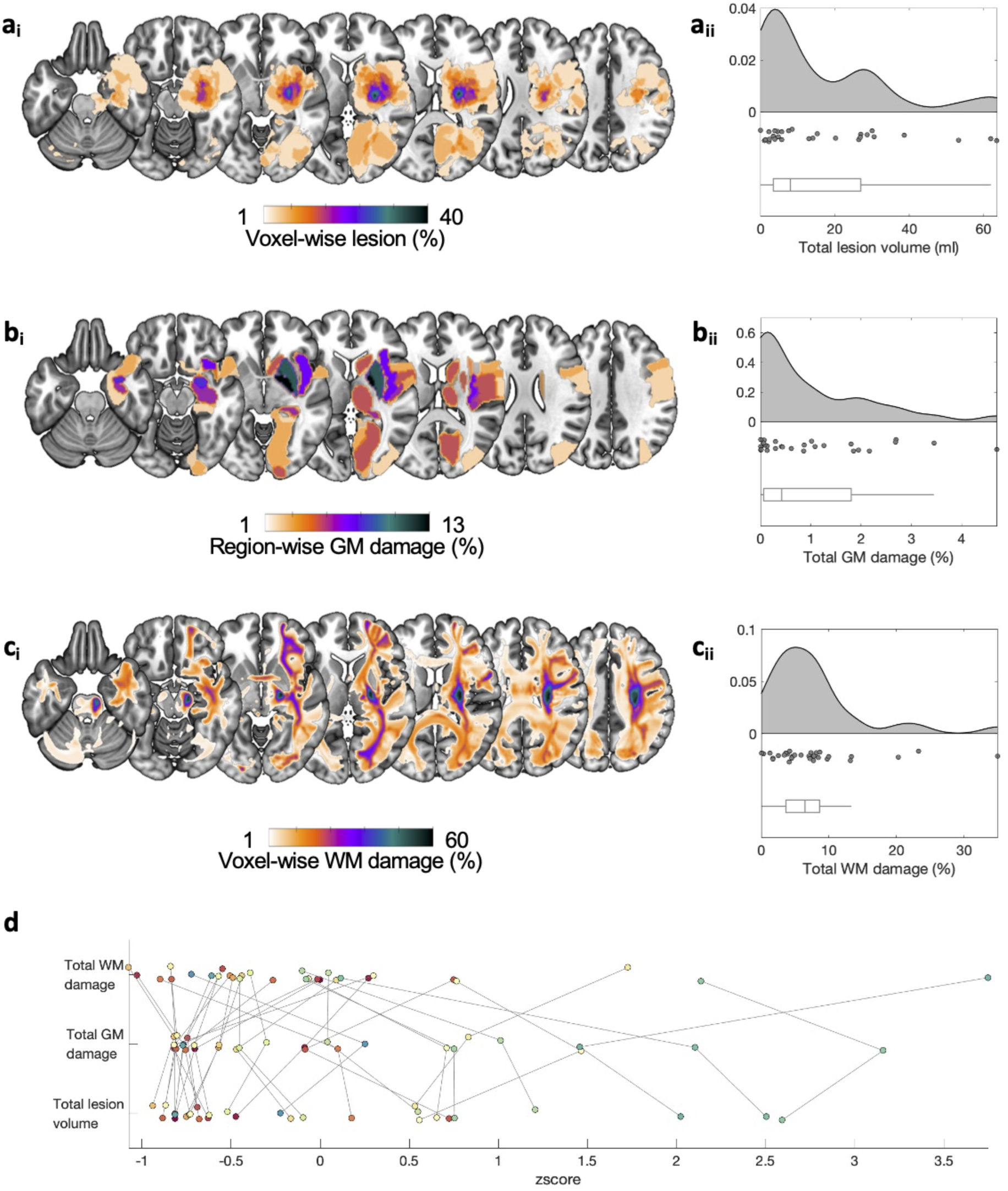
Lesion-related alterations in brain structure. **a_i_)** Group voxel-wise lesion map overlaid on an MNI template. Left hemispheric lesions were flipped. **a_ii_)** Total lesion volume as probability function (top), individual subjects, and boxplot (bottom). **b_i_)** Group region-wise map of GM damage overlaid on an MNI template. Left hemispheric lesions were flipped. **b_ii_)** Total GM damage as probability function (top), individual subjects, and boxplot (bottom). **c_i_)** Group voxel-wise map of WM damage overlaid on an MNI template. Left hemispheric lesions were flipped. **c_ii_)** Total WM damage as probability function (top), individual subjects, and boxplot (bottom). **d)** Normalised (zscore) total lesion volume, total GM damage, and total WM damage for each stroke survivor.

**Table 2.** Details of GM damage for the ten most damaged regions.

| Rank | Label | N of patients<br>with damage | M (%) ± SD (%) | Range<br>(%) |
| --- | --- | --- | --- | --- |
| 1 | Lenticular nucleus, Pallidum | 10/30 | 12.70 ± 26.40 | 0 – 98.04 |
| 2 | Lenticular nucleus, Putamen | 16/30 | 11.39 ± 20.29 | 0 – 82.07 |
| 3 | Amygdala | 8/30 | 8.49 ± 21.18 | 0 – 99.95 |
| 4 | Insula | 8/30 | 7.38 ± 15.76 | 0 – 63.21 |
| 5 | Hippocampus | 9/30 | 6.29 ± 15.88 | 0 – 56.99 |
| 6 | Heschl's gyrus | 6/30 | 5.53 ± 16.58 | 0 – 72.39 |
| 7 | Caudate nucleus | 12/30 | 5.36 ± 12.18 | 0 – 58.51 |
| 8 | Thalamus | 12/30 | 5.24 ± 17.33 | 0 – 92.08 |
| 9 | Calcarine fissure and<br>surrounding cortex | 5/30 | 4.90 ± 15.53 | 0 – 73.94 |
| 10 | Rolandic operculum | 8/30 | 4.86 ± 13.44 | 0 – 52.70 |

**Table 3.** Details of WM damage for the ten most damaged tracts.

| Rank | Label | N of patients<br>with damage | M (%) ± SD (%) | Range (%) |
| --- | --- | --- | --- | --- |
| 1 | Corticospinal Tract | 25/30 | 47.83 ± 39.04 | 0 – 100 |
| 2 | Parietopontine Tract | 26/30 | 39.84 ± 38.17 | 0 – 100 |
| 3 | Frontopontine<br>Tract | 24/30 | 31.08 ± 37.96 | 0 – 100 |
| 4 | Inferior Fronto Occipital<br>Fasciculus | 16/30 | 30.36 ± 40.74 | 0 – 100 |
| 5 | Arcuate Fasciculus | 14/30 | 28.52 ± 42.13 | 0 – 100 |
| 6 | Temporopontine Tract | 15/30 | 27.11 ± 40.05 | 0 – 100 |
| 7 | Anterior Commissure | 14/30 | 26.34 ± 39.73 | 0 – 99.91 |
| 8 | Extreme Capsule | 14/30 | 25.74 ± 39.75 | 0 – 100 |
| 9 | Occipitopontine Tract | 16/30 | 25.70 ± 36.39 | 0 – 100 |
| 10 | Superior Longitudinal Fasciculus | 13/30 | 19.53 ± 27.64 | 0 – 82.14 |

Next, we investigated stroke-related differences in brain function. *Passive movement – affected hand:* β-ERD (-0.35 to 0.30 s relative to passive movement) was lower in stroke survivors compared to HC in a large spatial cluster comprising 17 channels across contralateral (ipsilesional) and ipsilateral (contralesional) parietal and occipital channels (*threshold_cluster_forming_* = 3.5, *p_cluster_threshold_* = 0.01, *rank_cluster_* = 1, *T_cluster___sum_* = -404.13, *T_cluster_max_* = -5.33, *p_cluster_* < 0.001, **Fig. 3a_iii_**). β-ERS (0.95 to 2.25 s relative to passive movement) was lower in stroke survivors compared to HC in 5 channels across the contralateral (ipsilesional) sensorimotor area (*threshold_cluster_forming_* = 3.5, *p_cluster_threshold_* = 0.01, *rank_cluster_* = 2, *T_cluster___sum_* = 298.33, *T_cluster_max_* = 5.17, *p_cluster_* < 0.001, **Fig. 3a_iii_**). *Passive movement - unaffected hand:* No group difference was observed for β-ERD **(SI Fig. 1).** β-ERS was lower in stroke survivors compared to HC above contralateral (contralesional) and ipsilateral (ipsilesional) sensorimotor areas (**SI Fig. 1a, SI Table 1**). Stroke-related decreases in β-ERS are not specific to the affected hand. *Resting state:* There were no significant differences between HC and stroke survivors in resting state power at any frequency and any channel (**Fig. 3b**).

**Fig. 3.**
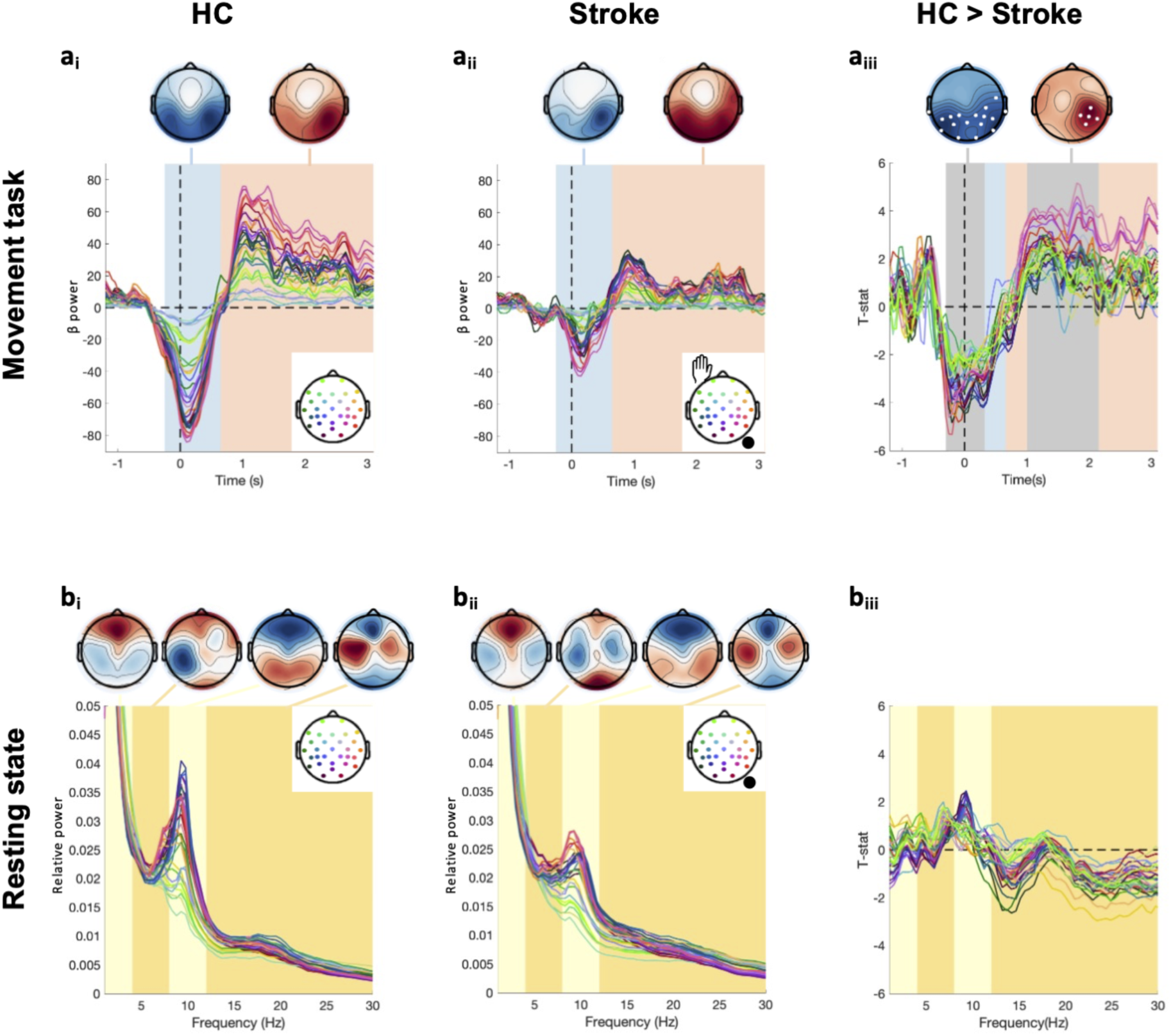
Brain function in acute stroke relative to healthy controls. **ai)** β power during passive movement for healthy controls. Mean power estimates as a function of time and space. The inlay illustrates the channel location colour coding. ERD and ERS time windows are highlighted in blue and red accordingly. Topographies show the spatial pattern of β power averaged over ERD and ERS time windows. Topographies are scaled from minus to plus the maximum absolute value of the plotted data. **aii)** Same as ai) for passive movement of the affected hand in stroke survivors. The inlay illustrates the channel location colour coding, the lesioned hemisphere (black circle) and the hand that was moved (here affected hand). **aiii)** t-values as a function of time and space for the contrast between healthy controls and acute stroke survivors. Statistical significance is assessed by time x channel cluster permutation testing. Significant times are indicated in by the grey area. Significant channels are highlighted in white in the topography. Topographies are scaled from plus and minus the maximum absolute value of the data plotted. **bi)** Resting state power for healthy controls. Mean power estimates as a function of frequency and space. The inlay illustrates the channel location colour coding. δ-, θ-, α-, β-frequency bands are highlighted in different shades of yellow. Topographies on the top show the spatial pattern of power averaged over each frequency band. Topographies are scaled from minus to plus the maximum absolute value of the plotted data. **bii)** Same as bi) for stroke survivors. The inlay illustrates the channel location colour coding, the lesioned hemisphere (black circle). **biii)** t-values as a function of frequency and space for the contrast between healthy controls and acute stroke survivors. Statistical significance is assessed by frequency x channel cluster permutation testing. No significant difference was found.

Next, we wanted to test whether stroke-related changes in brain structure and function are related. We found that stronger β-ERS induced by passive movement of the affected hand was related to lower lesion volume (*rho(28)* = -0.50, *p* = 0.005) and lower total GM damage (*rho(28)* = -0.61, *p* < 0.001), but not with total WM damage (*rho(28)* = -0.35, *p* = 0.057). Interestingly, a similar relationship was found between for β-ERS induced by passive movement of the unaffected hand with lesion volume (*rho(28)* = -0.42, *p* = 0.023) and total GM damage (*rho(28)* = -0.54, *p* = 0.002), but again not with total WM damage (*rho(28)* = - 0.08, *p* = 0.67). No significant relationships were observed between passive movement induced β-ERD (for either hand) and brain structure metrics (all *p’s* > 0.1).

### 3.3 Initial severity relates to brain structure but not brain function in acute stroke

We tested whether initial global or UL-specific severity were related to structural brain damage. Larger lesion volume was associated with greater initial global severity (*rho(28)* = 0.39, *p* < 0.05), but not with initial UL-specific severity (*p* > 0.1). Greater global initial severity was also associated with greater total GM damage (*rho(28)* = 0.40, *p* < 0.05, **Fig. 4aii**), while greater initial UL-specific severity was associated with to greater damage to the pallidum (*rho(28)* = 0.49, *p* < 0.05), putamen (*rho(28)* = 0.48, *p* < 0.01), and caudate (*rho(28)* = 0.47, *p* < 0.01, **Fig. 4bii**). Greater initial global severity was related to greater damage to the corticospinal (*rho(28)* = 0.40, *p* < 0.05), corticostriatal (*rho(28)* = 0.38, *p* < 0.05) and frontopontine tracts (*rho(28)* = 0.44, *p* < 0.05, **Fig. 4aiii**). Greater initial UL-specific severity was associated with greater damage to the corticospinal (*rho(28)* = 0.49, *p* < 0.01), corticothalamic (*rho(28)* = 0.41, *p* < 0.05), corticostriatal (*rho(28)* = 0.45, *p* < 0.05), parietopontine (*rho(28)* = 0.51, *p* < 0.01), and frontopontine tracts (*rho(28)* = 0.50, *p* < 0.01, **Fig. 4biii)**.

**Fig. 4.**
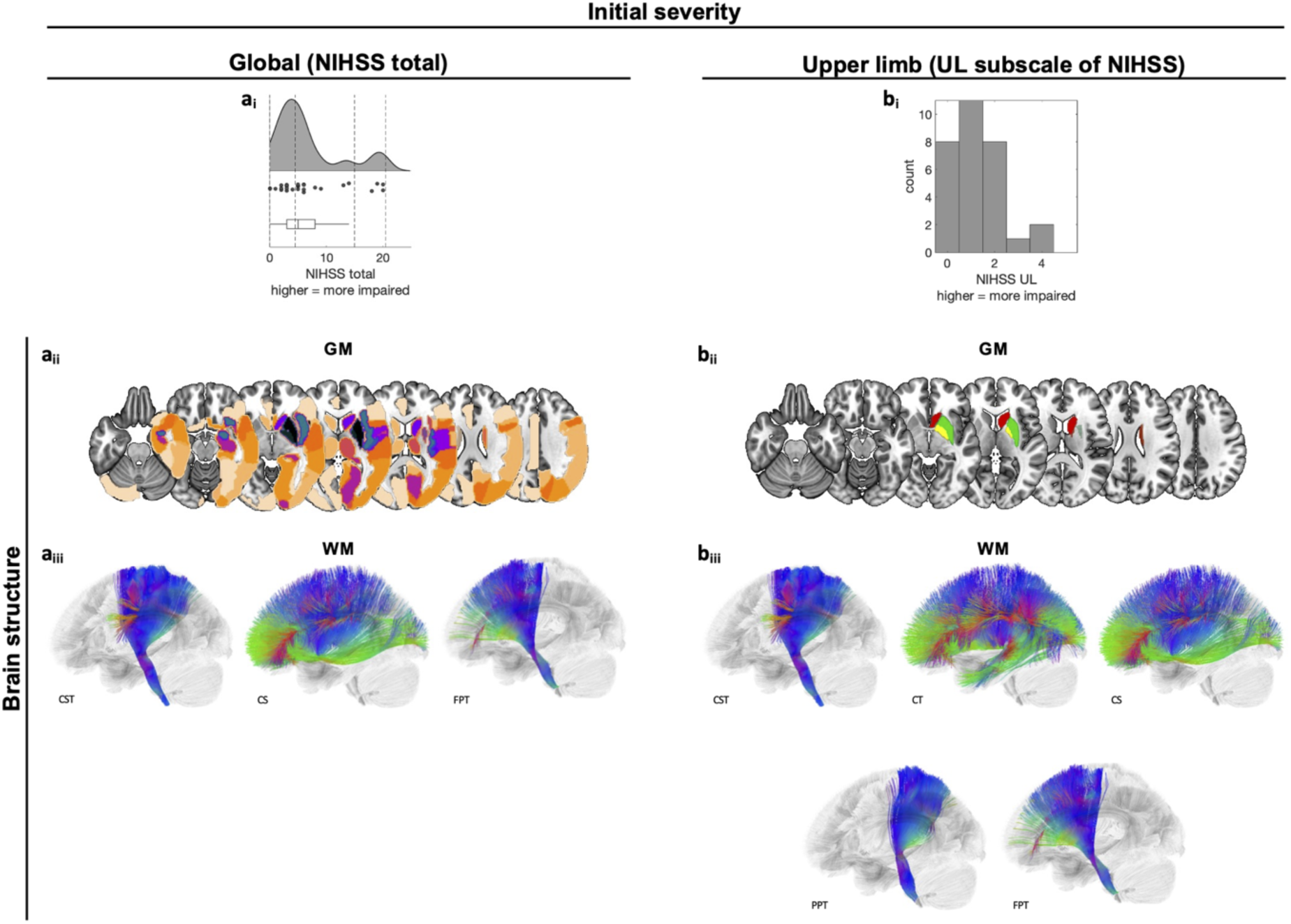
Relationship between brain structure and global and UL-specific initial severity. **ai)** Distribution of global initial severity (i.e., NIHSS total at hospital admission). Dashed lines represent different severity levels (minor stroke = 1-4, moderate stroke = 5-15, moderate to severe stroke = 16-20). **aii)** Global initial severity relates to total GM damage. Illustrated is the Group region-wise map of damage overlaid on an MNI template. Left hemispheric lesions were flipped. **aiii)** Global initial severity relates to greater WM damage of the corticospinal tract (CST), corticostriatal pathway (CS), and frontopontine tract (FPT). Tracts are colour-coded based on a standard red-green-blue (RGB) code applied to each (red for right-left, blue for dorsal-ventral, and green for anterior-posterior). **bi)** Distribution of UL-specific initial severity (i.e., UL subscale of NIHSS). **bii)** UL-specific severity relates to GM damage in the pallidum (yellow), putamen (green), caudate (red). **biii)** UL-specific initial severity relates to greater WM damage of the corticospinal tract (CST), corticothalamic pathway (CT), corticostriatal pathway (CS), parietopontine tract (PPT), and frontopontine tract (FPT). Tracts are colour-coded based on a standard red-green-blue (RGB) code applied to each (red for right-left, blue for dorsal-ventral, and green for anterior-posterior).

Regarding brain function, no significant relationships were found between global or UL-specific initial severity and resting state (any channel and any frequency) or β power induced by passive movement (any channel and any time point) (*threshold_cluster_forming_* = 3, *p_cluster_threshold_* = 0.01, **SI Fig. 2a, b**).

### 3.4 Subsequent recovery relates to brain function but not brain structure in acute stroke

We found no significant relationships between brain structure measures and global or UL-specific subsequent recovery after controlling for initial severity (all *p’s* > 0.1).

Regarding brain function, stronger β-ERD in the contralateral (ipsilesional) hemisphere, induced by passive movement of the affected hand, was associated with both global and UL-specific subsequent recovery (**Table 4**, **Fig. 5aii**, **Fig. 5bii**). This effect was specific to β-ERD induced by passive movement of the affected hand and not observed for the unaffected hand (*threshold_cluster_forming_* = 3, *p_cluster_threshold_* = 0.01, **SI Fig. 3**). For resting state activity, lower power in lower frequencies and higher power in higher frequencies were associated to both global and UL-specific subsequent recovery (**Table 4**, **Fig. 5aiii**, **Fig. 5biii**).

**Table 4.** Cluster statistic for significant clusters in contrasts examining the relationship between brain function and global and UL-specific subsequent recovery while accounting for initial severity. For all contrasts in this table applies: *threshold_cluster_forming_* = 3, *p_cluster_threshold_* = 0.01. Note the different N of subjects across comparisons, is because missing data in resting state recording, SIS, NISS. NIHSS is used as covariate in this analysis, to analyse subsequent recovery when accounting for initial severity.

| Index | Comparison | N of subjects | $rank_{cluster}$ | $T_{cluster\_sum}$ | $T_{cluster\_max}$ | $p_{cluster}$ | Time window | Frequency range | N of channels |
| --- | --- | --- | --- | --- | --- | --- | --- | --- | --- |
| 1 | Global (SIS) – Passive Movement Task | 26/30 | 1 | -92.68 | -4.01 | 0.030 | -0.15 – 0.20 s | NA | 10 |
| 2 |  | 26/30 | 2 | -78.42 | -3.94 | 0.036 | 0.20 – 0.35 s | NA | 13 |
| 3 | Global (SIS) – Resting State | 21/30 | 1 | 615.07 | 3.76 | 0.031 | NA | 10.25 – 22.46 Hz | 23 |
| 4 |  | 21/30 | 2 | -309.32 | -5.12 | 0.039 | NA | 2.44 – 6.35 Hz | 21 |
| 5 | UL subscale (SIS) – Passive Movement Task | 26/30 | 1 | -160.81 | -4.78 | 0.010 | -0.20 – 0.20 s | NA | 20 |
| 6 | UL subscale (SIS) – Resting State | 21/30 | 1 | 432.05 | 3.62 | 0.040 | NA | 14.16 – 31.25 | 20 |
| 7 |  | 21/30 | 2 | -312.32 | -5.07 | 0.042 | NA | 2.46 – 5.86 | 22 |

**Fig. 5.**
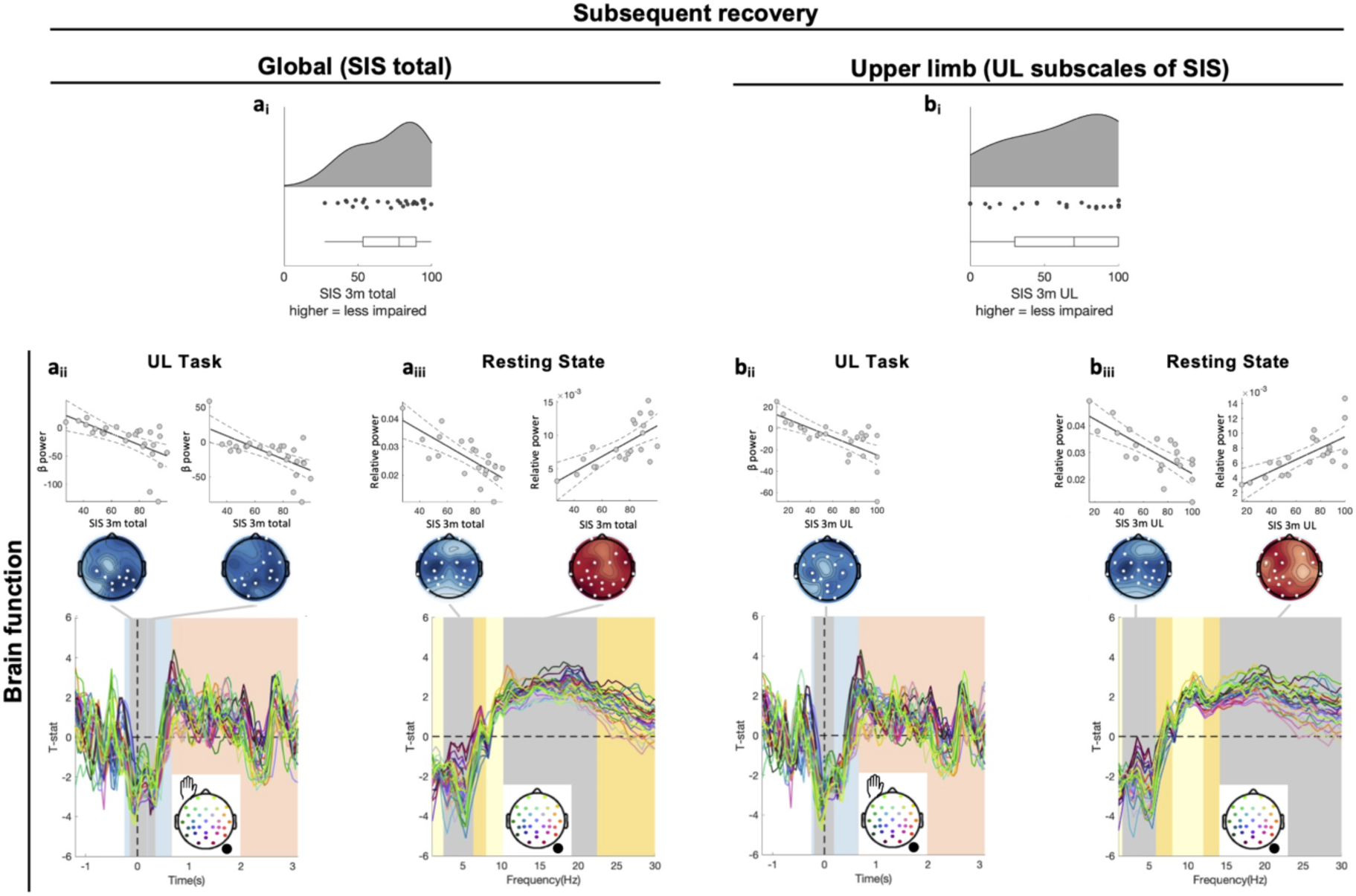
Relationship between brain function and global and UL-specific subsequent recovery while accounting for initial severity. **ai)** Distribution of global subsequent recovery (i.e., SIS total three months post-stroke). **aii)** (bottom) t-values as a function of time and space for the contrast correlating β power during passive movement of the affected hand and SIS total, while accounting for NIHSS total. The inlay illustrates the channel location colour coding, the lesioned hemisphere (black circle) and the hand that was moved (here affected hand). ERD and ERS time windows are highlighted in blue and red accordingly. Statistical significance is assessed by time x channel cluster permutation testing. Significant times are indicated by the grey area. Significant channels are highlighted in white in the topography. Topographies are scaled from plus and minus the maximum absolute value of the data plotted. Topographies are scaled from plus and minus the maximum absolute value of the data plotted. (top) Visual representation of the data in the significant cluster, i.e., the correlation between β power of the significant time x channel cluster and total SIS. **aiii)** (bottom) t-values as a function of frequency and space for the contrast correlating resting state power and SIS total, while accounting for NIHSS total. The inlay topography illustrates the channel location colour coding, the lesioned hemisphere (black circle). δ-, θ-, α-, β-frequency bands are highlighted in yellow. Statistical significance is assessed by frequency x channel cluster permutation testing. Significant frequencies are indicated by the grey area. Significant channels are highlighted in white in the topography. Topographies are scaled from plus and minus the maximum absolute value of the data plotted. (top) Visual representation of the data in the significant cluster, i.e., the correlation between resting state power of the significant time x channel cluster and total SIS. **bi)** Distribution of UL-specific subsequent recovery (i.e., SIS total three months post-stroke). **bii)** Same as ai) for UL-specific subsequent recovery. **biii)** Same as aii) for UL-specific subsequent recovery.

### 3.5 Combining clinical measures, brain structure, and brain function to predict subsequent recovery

While the previous analysis examined the direct relationship between either brain structure or brain function and initial severity or subsequent recovery, we next aimed to assess how relevant measures of brain structure and function contribute to predicting subsequent recovery. First, linear regression was used to test whether initial severity significantly predicted subsequent recovery. Global initial severity explained 42.9% of the variance in global subsequent recovery (*RMSE* = 57.1%, *F*(1,25) = 18.79, *p* < 0.001, see **SI Table 2** for coefficients), and the UL-specific initial severity explained 30.4% of the variance in UL-specific subsequent recovery (*RMSE* = 58.84, *F*(1,25) = 10.92, *p* = 003, see **SI Table 3** for coefficients). Next, adding brain structure (i.e., CST damage) as predictor to the model improved prediction (global subsequent recovery: *R^2^* = 59.1%, *RMSE* = 49.34, *F*(2,24) = 17.31, *p* < 0.001, see **SI Table 2** for coefficients; UL-specific subsequent recovery: *R^2^* = 46.8%, *RMSE* = 52.49, *F*(2,24) = 10.57, *p* < 0.001, see **SI Table 3** for coefficients). Then, adding brain function (i.e., task-related β-ERD) to the original model also improved prediction (global subsequent recovery: *R^2^* = 75.5%, *RMSE* = 38.19, *F*(2,24) = 36.93, *p* < 0.001, see **SI Table 2** for coefficients; UL-specific subsequent recovery: *R^2^* = 68.9%, *RMSE* = 40.12, *F*(2,24) = 26.62, *p* < 0.001, see **SI Table 3** for coefficients). Since the two β-ERD clusters identified in relation to global subsequent recovery showed strong collinearity (VIF = 4.95, 4.93), they were averaged before inclusion. Finally, a model combining initial severity, brain structure, and brain function provided the best prediction (global subsequent recovery: *R^2^* = 80.1%, *RMSE* = 35.13, *F*(3,23) = 30.87, *p* < 0.001, see **SI Table 2** for coefficients; UL-specific subsequent recovery: *R^2^* = 74.3%, *RMSE* = 37.30, *F*(3,23) = 22.12, *p* < 0.001, see **SI Table 3** for coefficients). The final model was confirmed via forward stepwise multivariate regression. For both global and UL-specific recovery, the full model — comprising initial severity, brain structure, and brain function — was retained.

## 4. Discussion

Here we asked how brain structure and function in acute stroke using standard imaging, clinical measures, and bedside EEG, relate to global and UL initial severity and subsequent recovery. Initial severity was linked to structural damage, but not to functional measures. In contrast, subsequent recovery was associated with brain function, but not structural damage. Nevertheless, the best prediction of recovery came from combining initial impairment, brain structure, and function, highlighting their complementary value.

### 4.1 Brain structure relates to initial severity but not subsequent recovery

We found that global initial severity was associated with lesion volume, total GM damage, and damage to specific WM tracts, including the CST, corticostriatal pathway, and frontopontine tract. No associations were observed with GM damage in individual regions. In contrast, UL-specific severity was linked to GM damage in the pallidum, putamen, and caudate, as well as damage to the CST, frontopontine, parietopontine, corticothalamic, and corticostriatal pathways.

The CST’s role in motor function explains its consistent association with both global and UL-specific severity. This aligns with prior work linking CST integrity (fractional anisotropy, lesion load, or fibre count) to acute motor impairment (NIHSS, FM-UE, ARAT, grip strength) (Aikio et al., 2021; Boccuni et al., 2019; Maraka et al., 2014; Saltão da Silva et al., 2022), and to somatosensory deficits (Boccuni et al., 2018). These relationships often persist beyond the acute stage (Schulz et al., 2012; Zhu et al., 2010). In addition to the CST, damage to the frontopontine and parietopontine tracts were also linked to initial severity. These tracts share anatomical proximity with the CST and are part of the corticopontine system (Prats-Galino et al., 2012). The observed associations with the corticothalamic and corticostriatal pathways implicate the cortico-basal ganglia-thalamocortical circuits (Mathai & Smith, 2011), which support motor planning and reinforcement learning (Graybiel et al., 1994). GM damage in the basal ganglia, in line with (Lin et al., 2023), may reflect disruption of these circuits. Initial severity was also associated with damage to the corticothalamic and corticostriatal pathways. These form part of the cortico-basal ganglia-thalamocortical circuits (Mathai & Smith, 2011) involved in motor planning and reinforcement learning (Graybiel et al., 1994). GM damage to the pallidum, putamen, and caudate, consistent with (Lin et al., 2023), may reflect disruption of these pathways.

We found no significant associations between structural brain damage and recovery. Though CST integrity has been linked to recovery (Feng et al., 2015; Saltão da Silva et al., 2022; Spampinato et al., 2017), many studies do not control for initial severity.

### 4.2 Brain function related to subsequent recovery but not initial severity

#### Brain function during task relates to subsequent recovery

While brain structure provides valuable insights, it is not sufficient on its own to predict recovery; incorporating measures of brain function is important for more accurate prediction of recovery trajectories. Consistent with prior studies, we observed significant differences in sensorimotor β-activity between stroke survivors in the acute stage post-stroke and HC when probing the sensorimotor system (Laaksonen et al., 2012; Parkkonen et al., 2017, 2018). Unlike earlier work focusing solely on post-movement β-ERS, we analysed the full β-ERD–ERS complex, revealing bilaterally reduced β-ERD and diminished ipsilesional β-ERS in stroke survivors. No association was found between β-activity and initial severity, but β-ERD was linked to later recovery.

Earlier MEG studies using tactile stimulation reported higher β-ERS correlating with lower initial severity and better recovery (Laaksonen et al., 2012; Parkkonen et al., 2017, 2018; Zich et al., 2025). Our study, using passive movement and bedside EEG, found a link between β-ERD and recovery. Differences in findings may stem from outcome measures (NIHSS vs. NHPT/BB) and stimulation type: (Laaksonen et al., 2012; Parkkonen et al., 2018) used tactile stimulation, while (Parkkonen et al., 2017, 2018) and this study used passive movement, known to elicit stronger β-ERS and comparable β-ERD in HC (Parkkonen et al., 2015). Additionally, while prior studies used MEG, we employed high-quality EEG, which, despite lower signal-to-noise ratio, shows strong cross-modality β-feature correlations (Illman et al., 2020). Sample variability (stroke type, lesion site, impairment severity, age, alertness) may also explain inter-study differences, especially given small acute stroke cohorts (Doric et al., 2025; Illman et al., 2021). Overall, early post-stroke β-activity may serve as a functional marker of motor recovery, though the distinct roles of β-ERD and ERS require further exploration.

#### Brain function during rest relates to subsequent recovery

We found no significant differences in resting-state spectral power (1–30 Hz) between acute stroke survivors and HC across EEG sensors. This aligns with MEG studies reporting no differences in β-peak amplitude at rest between hemispheres, time points, or groups (Aikio et al., 2021; Laaksonen et al., 2012; Parkkonen et al., 2017, 2018). Similarly, (Laaksonen et al., 2013) found no group or hemispheric differences in α/β power under eyes-open or eyes-closed conditions. In contrast, Wu and colleagues showed that δ/β power in specific clusters predicted initial severity in 25 stroke survivors 3–12 days post-stroke, with stronger δ power and weaker β power linked to worse outcomes (J. Wu et al., 2016).

Regarding recovery, lower resting δ/θ power and higher α/β power were associated with better global and upper limb recovery. These results align with previous findings linking elevated low-frequency power in acute stroke with poorer outcomes (Finnigan et al., 2004; Laaksonen et al., 2013). Such increases may reflect cortical deafferentation (Ball et al., 1977; Gloor et al., 1977), perilesional pathological activity (Kamada et al., 1997), or axonal sprouting (Carmichael & Chesselet, 2002). Additionally, our findings match results from a randomized trial where increased β power post-anodal tDCS was linked to improved lower limb function (Vimolratana et al., 2024). In contrast, (Laaksonen et al., 2013) found no link between α/β power and outcome, though they did not adjust for initial severity, possibly explaining the inconsistency.

Recovery-related power changes often showed bilateral topographies. Potential mechanisms for contralesional effects include mass effects (Macdonell et al., 1988; Newey et al., 2013), transcallosal diaschisis (Juhász et al., 1997), altered inhibition and excitability (Di Lazzaro et al., 2010, 2015; Liepert et al., 2000), and interhemispheric imbalance (Buetefisch, 2015).

### 4.3 Predicting subsequent motor recovery post-stroke

After examining the direct associations of brain structure and function with initial severity and subsequent recovery, we next assessed how these measures together predict recovery trajectories. Models that combined initial severity, brain structure, and brain function explained global and upper limb (UL)-specific recovery better than any single measure alone. Brain structure did not show a significant correlation with recovery on its own because correlation examines the relationship between two variables in isolation. However, in a regression model that includes other factors like initial severity and brain function, brain structure explains additional unique variance. This indicates it provides complementary information that enhances recovery prediction, even when its direct correlation with subsequent recovery is weak. The added value of combining brain function and structure has previously been demonstrated in predicting impairment (acute stage: (J. Wu et al., 2016), chronic stage (Stinear et al., 2007)), recovery (Tecchio et al., 2007; Zich et al., 2025), and therapy response in the subacute stage (Burke Quinlan et al., 2015). These findings suggest that stroke-related changes in brain function are not merely a consequence of structural damage; if they were, brain function would not account for variance in recovery beyond that explained by brain structure alone.

### 4.4 Limitations and future directions

There are a number of limitations with the present study, which are hopefully overcome by future research. While the sample size reflects the current landscape of research in acute stroke (Doric et al., 2025) it limits the conclusions we can draw from these results. Despite the challenges of acute stroke research more studies with larger, more diverse cohorts are needed to understand how stroke type, lesion location, and patient factors influence recovery. Our study focused on clinical tools (CT, MRI), standard assessments (e.g., NIHSS), and bedside-compatible EEG, while recovery was measured with the SIS—a validated, reliable, and remotely administered patient-reported outcome capturing disability and quality of life. However, additional measures of brain function, such as motor-evoked potentials from transcranial magnetic stimulation or functional network activity via fMRI, could provide further insight. Similarly, incorporating detailed motor assessments could further improve characterization of initial severity and subsequent recovery. Further exploration of the mechanisms underlying β-activity during task and rest will refine functional biomarkers. Developing standardized protocols that combine accessible bedside measures with advanced imaging can enhance individualized prognosis and support targeted therapies. These advances hold promise for improving stroke recovery outcomes and personalizing patient care.

## Acknowledgments

Thank you to all study participants. Thanks to the clinical team, the occupational therapist, physiotherapists, and radiologists at The National Hospital for Neurology and Neurosurgery (NHNN), Queen Square, for performing clinical assessment and structural brain imaging.

## Author contributions

CZ: Conceptualization, Methodology, Formal analysis, Investigation, Data Curation, Writing - Original Draft, Visualization, Project administration

LCM: Writing - Review & Editing, Project administration

AQ: Methodology, Software, Writing - Review & Editing

SB: Resources, Writing - Review & Editing

NSW: Conceptualization, Resources, Writing - Original Draft, Writing - Review & Editing, Funding acquisition

## Supplemental Information

**SI Fig. 1.**
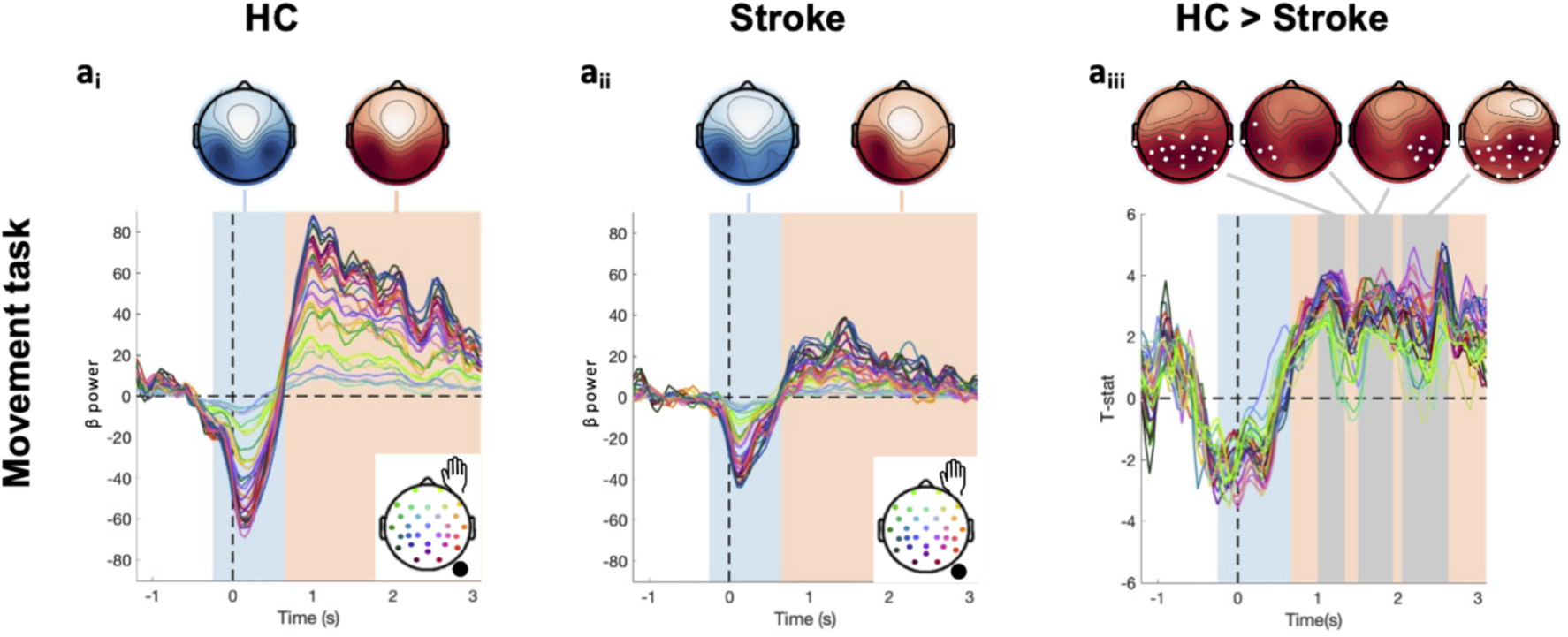
β power related to passive movement of the unaffected hand relative to healthy controls. **ai)** β power during passive movement for healthy controls. Mean power es9mates as a func9on of 9me and space. The inlay illustrates the channel loca9on colour coding. ERD and ERS 9me windows are highlighted in blue and red accordingly. Topographies show the spa9al pa@ern of β power averaged over ERD and ERS 9me windows. Topographies are scaled from minus to plus the maximum absolute value of the plo@ed data. **aii)** Same as ai) for passive movement of the unaffected hand in stroke survivors. The inlay illustrates the channel loca9on colour coding, the lesioned hemisphere (black circle) and the hand that was moved (here unaffected hand). **aiii)** t-values as a func9on of 9me and space for the contrast between healthy controls and acute stroke survivors. Sta9s9cal significance is assessed by 9me x channel cluster permuta9on tes9ng. Significant 9mes are indicated in by the grey area. Significant channels are highlighted in white in the topography. Topographies are scaled from plus and minus the maximum absolute value of the data plo@ed.

**SI Table 1.** Cluster statistic for significant clusters in contrasts examining the difference between β power induced by passive movement of the unaffected hand in stroke survivors and healthy controls (random hand). For all contrasts in this table applies: *threshold_cluster_forming_* = 3.5, *p_cluster_threshold_* = 0.01.

| Index | Comparison | $rank_{cluster}$ | $T_{cluster\_sum}$ | $T_{cluster\_max}$ | $p_{cluster}$ | Time window | N of channels |
| --- | --- | --- | --- | --- | --- | --- | --- |
| 1 | Healthy Controls > Acute | 1 | 301.73 | 5.07 | < 0.001 | 2.05 – 2.65 s | 19 |
| 2 |  | 2 | 229.06 | 4.20 | < 0.001 | 1.00 – 1.35 s | 17 |
| 3 |  | 3 | 82.69 | 4.27 | 0.002 | 1.5 – 1.9 s | 7 |
| 4 |  | 4 | 58.97 | 3.92 | 0.004 | 1.6 – 1.95 s | 5 |

**SI Fig. 2.**
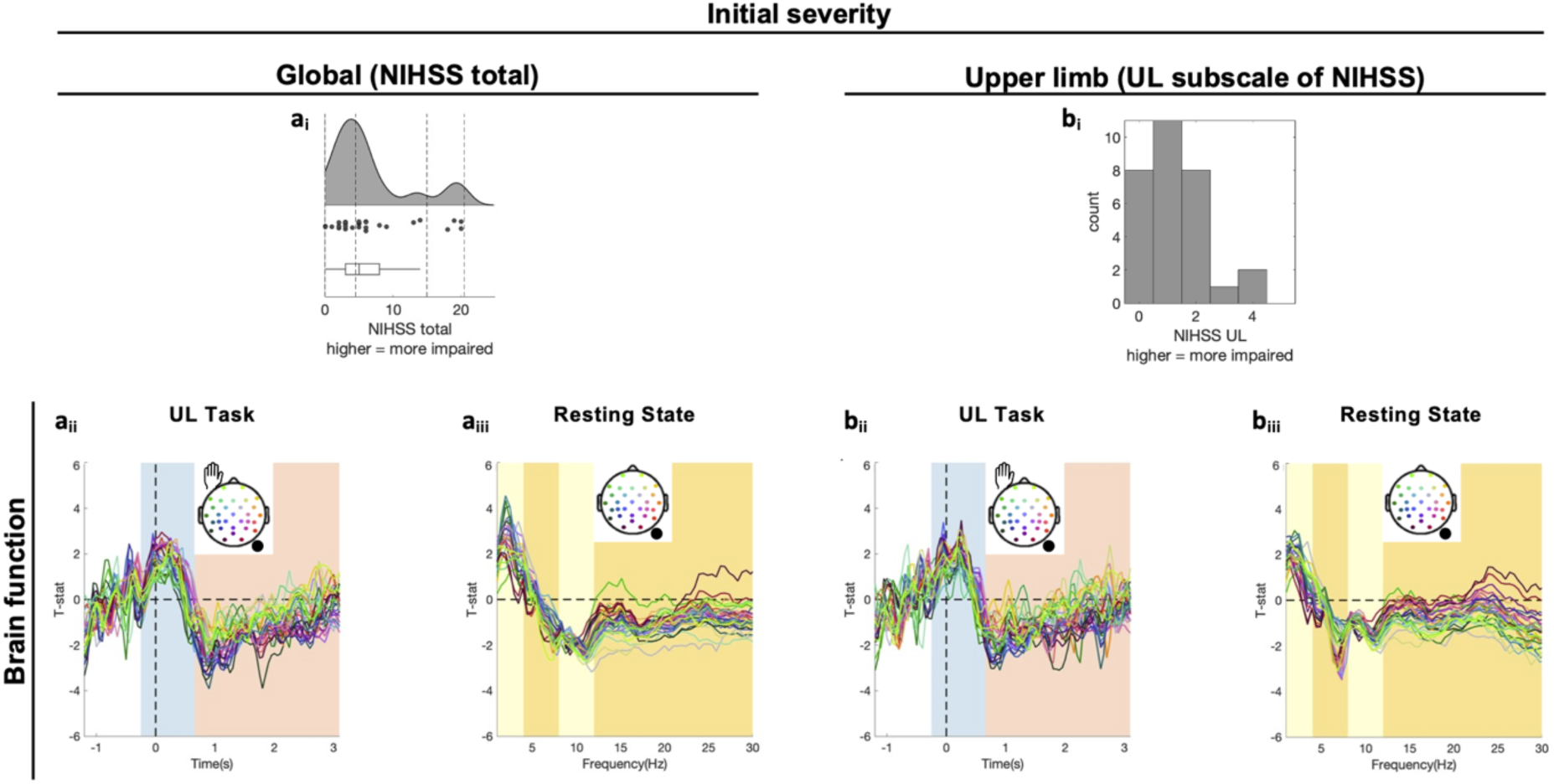
Relationship between brain function and global and UL-specific initial severity. **ai)** Global ini9al severity (i.e., NIHSS total from hospital admission) as probability function (top), individual subjects, and boxplot (bo@om). Dashed lines represent different severity levels (minor stroke = 1-4, moderate stroke = 5-15, moderate to severe stroke = 16-20). **aii)** (bo@om) t-values as a func9on of 9me and space for the contrast correlating β power during passive movement of the affected hand and SIS total, while accounting for NIHSS total. The inlay illustrates the channel loca9on colour coding, the lesioned hemisphere (black circle) and the hand that was moved (here affected hand). ERD and ERS 9me windows are highlighted in blue and red accordingly. Sta9s9cal significance is assessed by 9me x channel cluster permuta9on tes9ng. No significant cluster were found. **aiii)** (bo@om) t-values as a function of frequency and space for the contrast correla9ng res9ng state power and SIS total, while accounting for NIHSS total. The inlay topography illustrates the channel location colour coding, the lesioned hemisphere (black circle). δ, θ, α, β frequency bands are highlighted in yellow. Sta9s9cal significance is assessed by frequency x channel cluster permuta9on tes9ng. No significant cluster were found. **bi)** Distribution of UL-specific initial severity (i.e., UL subscale of NIHSS). **bii)** Same as ai) for UL-specific ini9al severity. **biii)** Same as aii) for UL-specific initial severity.

**SI Fig. 3.**
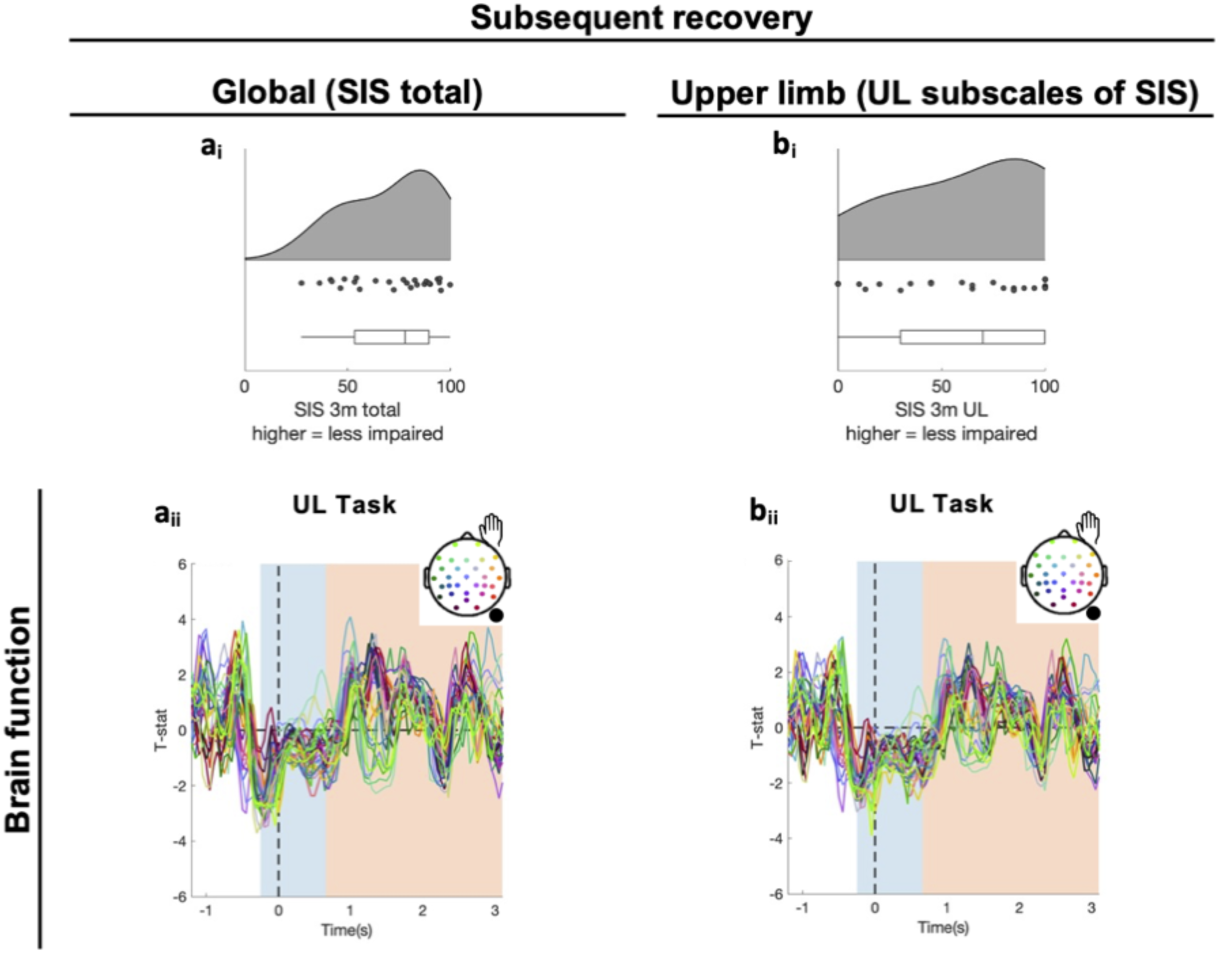
Relationship between β power induced by passive movement of the unaffected hand and global and UL-specific subsequent recovery while accounting for initial severity. **ai)** Global subsequent recovery (i.e., SIS total three months post stroke) as probability function (top), individual subjects, and boxplot (bo@om). **aii)** (bo@om) t-values as a func9on of 9me and space for the contrast correlating β power during passive movement of the unaffected hand and SIS total, while accounting for NIHSS total. The inlay illustrates the channel loca9on colour coding, the lesioned hemisphere (black circle) and the hand that was moved (here unaffected hand). ERD and ERS 9me windows are highlighted in blue and red accordingly. Sta9s9cal significance is assessed by 9me x channel cluster permuta9on tes9ng. No significant cluster were found. **bi)** UL-specific subsequent recovery (i.e., SIS total three months post stroke) as probability function (top), individual subjects, and boxplot (bo@om). **bii)** Same as ai) for UL-specific subsequent recovery.

**SI Table 2.** The coefficient table of the regression analysis for global subsequent recovery.

| Index | Variable | Unstand. coeff. |  | Sta. coeff. | *t* | *p* | 95% CI |  |
| --- | --- | --- | --- | --- | --- | --- | --- | --- |
| | | $\beta$ | *SE* | $\beta$ | | | Lower | Upper |
| 1 | NIHSS | 5.48 | 1.26 | 1.58 | 4.34 | <0.001 | 2.88 | 9.08 |
| 2 | NIHSS | 2.61 | 1.44 | 0.75 | 1.82 | 0.081 | -0.35 | 5.57 |
|  | CST | 0.65 | 0.21 | 1.12 | 3.08 | 0.005 | 0.21 | 1.09 |
| 3 | NIHSS | 4.63 | 0.86 | 1.34 | 5.39 | <0.001 | 2.85 | 6.40 |
| | $\beta$ activity | -1.22 | 0.22 | -1.71 | -5.64 | <0.001 | -1.66 | -0.77 |
| 4 | NIHSS | 3.11 | 1.03 | 0.90 | 3.03 | 0.006 | 0.98 | 5.23 |
|  | CST | 0.37 | 0.16 | 0.68 | 2.31 | 0.03 | 0.04 | 0.71 |
| | $\beta$ activity | -1.05 | 0.21 | -1.47 | -4.93 | <0.01 | -1.48 | -0.61 |

**SI Table 3.** The coefficient table of the regression analysis for UL-specific subsequent recovery.

| Index | Variable | Unstand. coeff. |  | Sta. coeff. | *t* | *p* | 95% CI |  |
| --- | --- | --- | --- | --- | --- | --- | --- | --- |
| | | $\beta$ | *SE* | $\beta$ | | | Lower | Upper |
| 1 | NIHSS | 22.91 | 6.93 | 0.87 | 3.31 | 0.003 | 8.63 | 37.19 |
| 2 | NIHSS | 6.94 | 8.53 | 0.26 | 0.81 | 0.42 | -10.66 | 24.53 |
|  | CST | 0.64 | 0.24 | 0.83 | 2.72 | 0.012 | 0.156 | 1.13 |
| 3 | NIHSS | 20.393 | 4.75 | 4.75 | 4.29 | <0.001 | 10.59 | 30.20 |
| | $\beta$ activity | -2.11 | 0.39 | 0.39 | -5.46 | <0.001 | -2.91 | -1.31 |
| 4 | NIHSS | 11.15 | 6.12 | 0.42 | 1.82 | 0.081 | -1.51 | 23.80 |
|  | CST | 0.38 | 0.18 | 0.50 | 2.18 | 0.039 | 0.02 | 0.75 |
| | $\beta$ activity | -1.87 | 0.38 | -1.15 | -4.95 | <0.01 | -2.65 | -1.09 |

